# Interleukin-6 in COVID-19: A Systematic Review and Meta-Analysis

**DOI:** 10.1101/2020.03.30.20048058

**Authors:** Eric A. Coomes, Hourmazd Haghbayan

## Abstract

**Purpose:** Coronaviruses may activate dysregulated host immune responses. As exploratory studies have suggested that interleukin-6 (IL-6) levels are elevated in cases of complicated COVID-19 and that the anti-IL-6 biologic tocilizumab may be beneficial, we undertook a systematic review and meta-analysis to assess the evidence in this field.

**Methods:** We systematically searched MEDLINE and EMBASE for studies investigating the immunological response in COVID-19 or its treatment with tocilizumab; additional grey literature searches were undertaken. Meta-analysis was undertaken using random effects models.

**Results:** Eight published studies, three pre-prints, and five registered trials were eligible. Meta-analysis of mean IL-6 concentrations demonstrated 2.9-fold higher levels in patients with complicated COVID-19 compared with patients with non-complicated disease (six studies; n=1302; 95%CI, 1.17-7.19; *I_2_*=100%). A single non-randomized, single-arm study assessed tocilizumab in patients with severe COVID-19, demonstrating decreased oxygen requirements, resolution of radiographic abnormalities, and clinical improvement. No adverse events or deaths were observed.

**Conclusions:** In patients with COVID-19, IL-6 levels are significantly elevated and associated with adverse clinical outcomes. While inhibition of IL-6 with tocilizumab appears to be efficacious and safe in preliminary investigation, the results of several ongoing clinical trials should be awaited to better define the role of tocilizumab in COVID-19 prior to routine clinical application.

**PROSPERO Registration:** CRD42020175879

## INTRODUCTION

A novel coronavirus, severe acute respiratory syndrome-coronavirus 2 (SARS-CoV-2), emerged in December 2019 from Wuhan, China.^1,2^ Causing a febrile respiratory illness known as coronavirus disease 2019 (COVID-19), this is the third zoonotic coronavirus to infect humans in the past two decades.^3^ Compared to its predecessors, COVID-19 has demonstrated rapid capacity for dissemination, having already infected over 500,000 patients worldwide.^4^ Such transmission has been fuelled by the high intrinsic reproductive number of 2-2.5,^5-7^ burgeoning community transmission,^8-10^ and potential occult transmission during the pre-symptomatic incubation period.^11-13^ In China, nearly one-fifth of infected patients experience severe or critical illness,^14^ with an overall 2.3% case fatality rate and up to 6.1% of patients experiencing severe complications.^15^ As preventative vaccines and effective antivirals remain unavailable, host-directed therapeutics employing existing immunomodulatory agents must be explored.^16,17^

Coronaviruses have been observed to activate excessive and dysregulated host immune responses which may contribute to the development of ARDS.^18,19^ Autopsy analyses of patients with COVID-19 complicated by ARDS have revealed hyperactivation of cytotoxic T-cells, with high concentrations of cytotoxic granules.^20^ Reports describing the immunological profile of critically ill patients with COVID-19 have suggested hyperactivation of the humoral immune pathway – including interleukin (IL)-6 – as a critical mediator for respiratory failure, shock, and multiorgan dysfunction. Given the potential for the development of cytokine release syndrome (CRS) as pathologic underpinning for disease progression of severe COVID-19 infection, this dysregulation of host immune responses may be an important target for therapeutics.

The biologic agent tocilizumab is a humanized monoclonal antibody targeting IL-6 receptors, blocking downstream pro-inflammatory effects of IL-6. It is approved by the Food and Drug Association for CRS induced by chimeric antigen receptor-T cell therapies and immune-mediated rheumatic diseases.^21,22^ Exploratory studies have suggested that IL-6 inhibition may be beneficial in patients with severe COVID-19 and, despite the absence of definitive evidence, very high drug cost, and unclear potential for harm, it has already been incorporated into COVID-19 management guidelines generated in China and Italy.^23,24^ This may thus incite clinical use without definitive evidence supporting its safety in COVID-19. We therefore designed a systematic review and meta-analysis to assess the evidence describing IL-6 response in patients with COVID-19 and the existing and planned studies investigating the efficacy of tocilizumab to guide patient management, clarify guideline development, and inform future trials in this field.

## METHODS

### Design

We undertook a systematic review and meta-analysis investigating IL-6 dysregulation in patients diagnosed with COVID-19, and the potential role of IL-6 suppression with the administration of tocilizumab. Articles eligible for inclusion were observational cohort, case-control, or randomized controlled trials (RCTs) characterizing either serum IL-6 dynamics or therapeutic response to tocilizumab in adult or pediatric patients diagnosed with COVID-19. This systematic review was undertaken with methodology in accordance with *Cochrane Handbook*,^25^ and reporting consistent with the Preferred Reporting Items for Systematic Reviews and Meta-Analyses (PRISMA).^26^ An *a priori* protocol was designed and registered (PROSPERO identification: CRD42020175879).

### Search Strategy

We designed a high sensitivity search strategy combining free text and keyword search term synonym clusters for COVID-19, combined with clusters for either IL-6 or tocilizumab. We then systematically searched for published articles in Ovid Medline Embase, pre-publication manuscripts in pre-print servers (Biorxiv, Medrxiv, and Chinxiv), and Google Scholar, and for planned trials in clinical trial registries (clinicaltrials.gov and the Chinese Clinical Trial Registry). All searches spanned January 1, 2019 to March 15, 2020; no exclusions were made for language, disease severity, or outcomes reported. We then conducted an expanded Ovid Medline and Embase database search from January 1, 2020 to March 15, 2020 for all published cohort studies reporting COVID-19 patient characteristics and outcomes (see *Appendix* for full search strategy) to ensure all studies reporting IL-6 levels in COVID-19 were identified. Reference lists of all included articles were also reviewed for potential eligibility of citations.

### Study Selection and Data Extraction

Two reviewers (E.A.C. and H.H.) independently undertook two-step selection, with studies screened via titles and abstracts followed by full-text review. Studies were included if they were RCTs, observational cohorts, or case-control in design, describing two or more patients diagnosed with COVID-19, and reported either measures of cytokine levels (with a focus on IL-6) or tocilizumab administration.

Data extraction was undertaken in duplicate (E.A.C. and H.H.) via standardized data extraction tables. Data were extracted from article text, tables, and graphs (employing figure analysis tools to quantitatively estimate curves for data). Data were collected for study design and setting, patient demographics, disease characteristics, levels of immune markers and indicators of systemic inflammation (inflammatory markers and cytokine levels), immunomodulatory agents administered (corticosteroids and intravenous immunoglobulin (IVIg)), and outcomes consistent with complicated infection (hospitalization, intensive care unit (ICU) admission, ARDS, invasive mechanical ventilation, renal replacement therapy, severe disease on clinical scoring tools (such as the Chinese New Coronavirus Pneumonia Prevention and Control Program or any others), or death). Conflicts were resolved by consensus discussion.

### Statistical Analysis

Count data and nominal variables are presented as proportions with percentages while continuous data are presented as means and standard deviations (SDs), or medians and interquartile ranges (IQR) or range. Measures of association relating clinical characteristics or receipt of tocilizumab with downstream clinical outcomes are presented in both unadjusted and adjusted forms, as availability of data permitted.

Results are described and summarized quantitatively and semi-qualitatively; for data deemed adequately homogenous in terms of patient characteristics, interventions, and clinical outcomes, meta-analysis was undertaken using random effects models. For statistical homogeneity, medians and IQRs were converted to means with SDs to maximize the number of studies eligible for meta-analysis.^27^ For such continuous data, we computed ratio of means (RoM) for each study and undertook meta-analysis via generic inverse variance methods (DerSimonian and Laird) to produce pooled measures of association, corresponding 95% confidence intervals (95%CI), and forest plots.^25,28,29^ Pre-specified subgroup analyses were conducted in regard to individual sub-definitions of complicated disease (as defined by primary studies investigators).

A pre-specified alpha of 0.05 was used for all statistical tests and confidence intervals; statistical heterogeneity was assessed by the *I*^*2*^ statistic. Data analysis was undertaken utilizing Microsoft Excel version 16.35 (Microsoft, Redmond, United States, 2020) and Review Manager version 5.3.5 (Cochrane Collaboration, Copenhagen, Denmark, 2014).

### Risk of Bias Assessment

Two reviewers (E.A.C. and H.H.) independently rated all included studies for risk of bias. The updated Quality in Prognostic Studies (QUIPS) tool was employed for cohort studies associating IL-6 levels with disease severity and the Risk of Bias in Non-Randomized Studies of Interventions (ROBINS-I) tool for non-randomized therapeutic studies of tocilizumab.^30-32^

## RESULTS

Following removal of duplicates, our database search identified 1,219 unique citations, of which 112 articles were assessed via full text and eight studies were eligible for inclusion (Figure 1). An additional three articles were identified via pre-print server searches; five planned or in-process clinical trials were identified via clinical trial registry searches. A total of 16 articles were therefore eligible for inclusion, with eleven (n=1819) contributing to qualitative synthesis and six (n=1302) undergoing quantitative synthesis (meta-analysis) (Figure 1).

**Figure 1.**
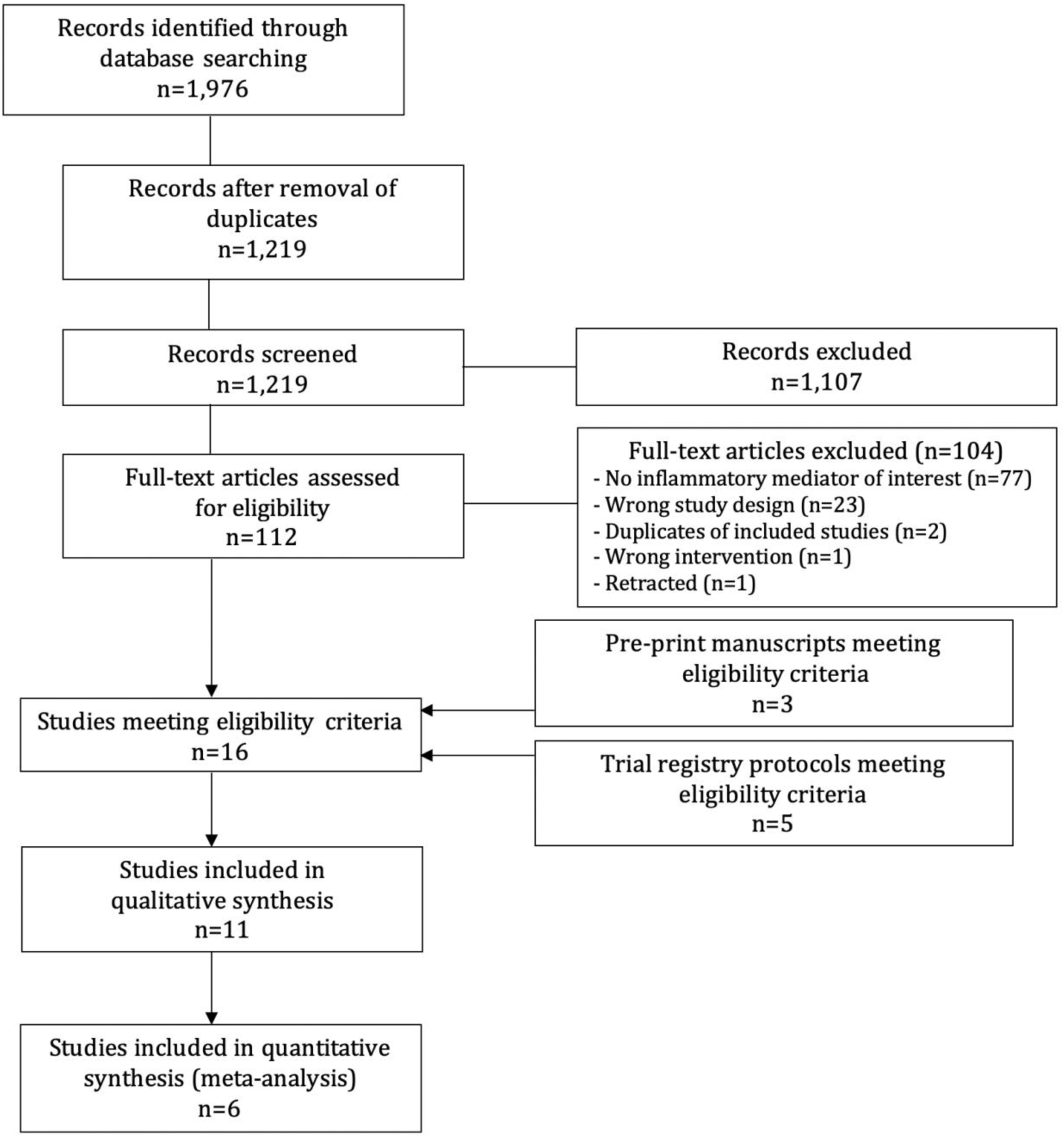
Study Selection Flow.

Individual study characteristics and patient demographics are presented in Table 1, and inflammatory markers, therapeutic interventions, and disease complications are presented in Table 2. The summary of registered clinical trials planned or underway investigating tocilizumab treatment for COVID-19 is presented in Table 3.

**Table 1.** Methodological and Patient Characteristics of the Studies Eligible for Inclusion

| Study (Year) | Location | Setting | Design | No of Participants (n) | Age, years (mean +/- SD) | Sex, M/F | Disease Severity<sup>1</sup> (n, %) | ICU admission, n (%) | ARDS, n (%) | Invasive Mechanical ventilation, n (%) |
| --- | --- | --- | --- | --- | --- | --- | --- | --- | --- | --- |
| **Chen et al. 2020a**<sup>33</sup> | Wuhan, China | Hospital inpatients | Prospective cohort; single centre | 29 COVID-19 patients | Median 56 (range 26-79) | 72%/28% | Mild (15; 52%)<br>Severe (9; 31%)<br>Critical (5; 17%) | NR | NR | NR |
| **Chen et al. 2020b**<sup>34</sup> | Wuhan, China | Hospital Inpatients | Retrospective cohort; single centre | 99 COVID-19 patients | 55.5 ± 13.1 | 68%/32% | NR | 23 (23%) | 17 (17%) | 4 (4%) |
| **Diao et al. 2020**<sup>40</sup> | Wuhan, China | Hospital inpatients | Retrospective cohort, multicentre | 552 COVID-19<sup>2</sup> patients; 40 healthy controls | NR | NR | NR | 20 (4%)<sup>1</sup><br>N=499 | NR | NR |
| **Huang et al. 2020a**<sup>36</sup> | Wuhan, China | Hospital inpatients | Prospective cohort; single centre | 41 COVID-19<sup>2</sup> patients; 4 controls | Median 49 (IQR 41-58) | 73%/27% | NR | 13 (32%) | 12 (29%) | 4 (10%) |
| **Huang et al. 2020b**<sup>35</sup> | Wuhan, China | Hospital inpatients | Retrospective cohort; single centre | 34 COVID-19 patients | 56 ± 17.1 | 41%/59% | NR | 8 (24%) | NR | 3 (9%) |
| **Liu et al. 2020**<sup>37</sup> | Wuhan, China | Hospital inpatients | Retrospective cohort; single centre | 80 COVID-19 patients | Median 53 (range 26-86) | 43%/57% | Mild (11, 14%)<br>Severe (69, 86%) | 3 (4%) | 7 (9%) | 2 (2.5%) |
| **Qin et al. 2020**<sup>38</sup> | Wuhan, China | Hospital inpatients | Retrospective cohort; single centre | 452 COVID-19 patients | Median 58 (IQR 47-67) | 52%/48% | Severe (286, 63%) | NR | NR | NR |
| **Ruan et al. 2020**<sup>39</sup> | Wuhan, China | Hospital inpatients | Retrospective cohort; multicentre | 150 COVID-19 patients | Died (68 patients):<br>Median 67 (range 15-81)<br>Discharged (82 patients)<br>Median 50 (range 44-81) | 68%/32% | NR | 41 (27%) | 62 (41%) | 25 (17%) |
| **Wu et al. 2020**<sup>41</sup> | Wuhan, China | Hospital inpatients | Retrospective cohort, multicentre | 201 COVID-19 patients | Median 51 (IQR 43-60) | 64%/36% | NR | 53 (26%) | 84 (42%) | 6 (3%) |
| **Xu et al. 2020**<sup>43</sup> | Anhui, China | Hospital inpatients | Non-randomized open-label clinical trial of tocilizumab for severe COVID-19 | 21 COVID-19 patients | 56.8 ± 16.5 | 86%/14% | Severe: (17, 81%)<br>Critical (4, 19%) | NR | NR | 2 (10%) |
| **Zhu et al. 2020**<sup>42</sup> | Anhui, China | Emergency department, patients under investigation | Retrospective cohort, multicentre | 32 COVID-19<sub>2</sub> patients; 84 negative cases | Median 46 (IQR 35-52) | 47%/53% | Mild (32, 100%) | NR | NR | NR |

1. Data refer only to patients diagnosed with COVID-19. 2. As per the Chinese New Coronavirus Pneumonia Prevention and Control Program score.

**Table 2.** Cytokine Levels and Clinical Outcomes in Patients with COVID-19

| Study (Year) | IL-6, pg/mL (mean ± SD) | Lymphocyte cells x 10<sup>9</sup> (mean ± SD) | Ferritin mcg/L (mean ± SD) | CRP, mg/L (mean ± SD) | ESR, mm/h (mean ± SD) | Corticosteroid Therapy, n (%) | IVIg, n (%) | Hospitalization, n (%) | Death, n (%) | Renal Replacement Therapy, n (%) |
| --- | --- | --- | --- | --- | --- | --- | --- | --- | --- | --- |
| **Chen et al. 2020a**<sup>33</sup> | Severe/critical: 72 ± 12<br>Non-severe: 34 ± 7 | “Decreased” <1.0 (69%) | NR | “Increased” >5 (93%) | NR | NR | NR | 29 (100%) | 2 (7%) | NR |
| **Chen et al. 2020b**<sup>34</sup> | Median 7.9 (IQR 6.1-10.6) | 0.9 ± 0.5 | 808.7 ± 490.7 | 51.4 ± 41.8 | 49.9 ± 2 3.4 | 19 (19%) | 27 (27%) | 99 (100%) | 11 (11%) | 9 (9%) |
| **Diao et al. 2020**<sup>40</sup> | ICU: 186 ± 283<sub>1</sub><br>Non-ICU: 51 ± 74<sub>1</sub> | NR | NR | NR | NR | NR | NR | NR | NR | NR |
| **Huang et al. 2020a**<sup>36</sup> | ICU: median 6.1 (IQR 1.8-37.7)<sub>1</sub><br>Non-ICU: median 5 (IQR 0-11.2) | Median 0.8 IQR 0.6–1.1 | NR | NR | NR | 9 (22%) | NR | 41 (100%) | 6 (15%) | 3 (7%) |
| **Huang et al. 2020b**<sup>35</sup> | “Increased” (9/9 tested patients) | Decreased (50%) | NR | NR | Increased (59.1%) | 21 (62%) | NR | 33 (97.1%) | NR | NR |
| **Liu et al. 2020**<sup>37</sup> | Severe: median 36.5 (IQR 30.8-42)<br>Non-severe: median 2.4 (IQR 2.1-2.9) | “Decreased” <1.5 (75%) | 690.2 ± 864.3 | “Increased” >10 (75%) | 40.6±27.2 | 29 (36%) | 36 (45%) | 80 (100%) | 0 (0%) | 0 (0%) |
| **Qin et al. 2020**<sup>38</sup> | Severe: median 25.5 (IQR 9.5-54.5)<br>Non-severe: median 13.3 (IQR 3.9-41.1) | Median 0.9 (IQR 0.6-1.2) | Median 662.4 (IQR 380.9-1311.9) | Median 44.1 (IQR 15.5-93.5) | Median 31.5 IQR 17.0-58.0 | NR | NR | 452 (100%) | NR | NR |
| **Ruan et al. 2020**<sup>39</sup> | 8.9 ± 6.7 | 1.0 ± 1.6 | 923.9 ± 949.7 | 76.0 ± 94.0 | NR | 53 (35%) | NR | 150 (100%) | 68 (45%) | 5 (3%) |
| **Wu et al.**<br>**2020<sup>41</sup>** | ARDS: median 7.4<br>(IQR 5.6-10.9)<br>No ARDS: median<br>6.3 (IQR 5.4-7.8) | Median 0.91<br>(IQR 0.61-1.29) | Median 594.0<br>(IQR 315.7-1266.2) | Median 42.4<br>(IQR 14.2-92.7) | Median 49.3<br>IQR 40.0-66.9 | 62 (31%) | NR | 201 (100%) | 44 (22%) | NR |
| **Xu et al.**<br>**2020<sup>43</sup>** | 132.38 ± 278.53 | Lymphocyte<br>percentage:<br>15.52% ± 8.89 | NR | 75.06 ± 66.8 | NR | 21 (100%) | NR | 21 (100%) | 0 (0%) | NR |
| **Zhu et al.**<br>**2020<sup>42</sup>** | “Increased” in 7/32<br>(22%)<sub>1</sub> | 1.1 ± 0.6<sub>1</sub> | NR | 20.7 ± 24.0<sub>1</sub> | 42.4 ± 33.6<sub>1</sub> | NR | NR | NR | NR | NR |

1. Data refer to patients with COVID-19.

**Table 3.** Summary of Registered Clinical Trials Investigating the Therapeutic Effect of Anti-IL-6 Inhibition for the Treatment of COVID-19

| ID | Status | Design | Country | Population | Intervention | Comparator | Primary Outcome |
| --- | --- | --- | --- | --- | --- | --- | --- |
| NCT04310228<sup>53</sup> | Recruiting | Multi-centre randomized controlled trial | China | COVID-19 pneumonia (n = 150) | Favipiravir plus Tocilizumab | Favipiravir monotherapy OR Tocilizumab monotherapy | Cure rate |
| NCT04306705<sup>52</sup> | Recruiting | Retrospective Cohort | China | Severe COVID-19 pneumonia with elevated IL-6 levels (n = 120) | Tocilizumab | 1) Continuous renal replacement therapy<br>2) Conventional therapy | Normalization of Fever and Oxygen Saturation within 14 days of treatment |
| ChiCTR2000029765<sup>49</sup> | Recruiting | Multi-centre randomized controlled trial | China | COVID-19 pneumonia with elevated IL-6 levels (n = 188) | Tocilizumab | Conventional therapy | Cure rate |
| ChiCTR2000030196<sup>50</sup> | Not yet recruiting | Multi-centre non-randomized open-label intervention | China | Severe COVID-19 pneumonia with elevated IL-6 levels, and grade 2-3 cytokine-release syndrome (n = 60) | Tocilizumab | N/A | Resolution of cytokine release syndrome |
| ChiCTR2000030442<sup>51</sup> | Cancelled by Investigator | Single-Centre Non-randomized intervention | China | Severe COVID-19 pneumonia (n=100) | Tocilizumab, intravenous immunoglobulin, and CRRT | N/A | Duration of hospitalization |

### Interleukin-6 and COVID-19

Ten cohort studies (n=1798) described the immunological response to SARS-CoV-2 in patients diagnosed with COVID-19; mean age was 54.8±14.4 and 42% were female. All studies were set in China and all but one exclusively recruited hospital inpatients. Of studies reporting the use of immunomodulatory therapies, corticosteroids were the most commonly administered agents and were received by 32% of patients. In studies reporting survival, global mortality was 22% among patients diagnosed with COVID-19 (Tables 1 and 2).

Overall, elevations in IL-6 levels amongst patients with COVID-19 were identified in all included studies.^33-42^ Multiple studies specifically identified higher levels of IL-6 amongst patients with more severe (complicated) disease.^33,37-40^ Descriptions of other inflammatory markers, including IL2R and ferritin, are contained in the Supplementary Appendix. A total of six studies (n=1302) compared IL-6 levels in patients with complicated disease (patients with ARDS, requiring ICU admission, or determined to have either “severe” or “critical” presentations as per the Chinese New Coronavirus Pneumonia Prevention and Control Program score) with non-complicated disease (none of the above criteria present) and were included in meta-analysis. Compared to patients with non-complicated disease, IL-6 levels in those with complicated COVID-19 were 2.90-fold higher (six studies; n=1302 patients; 95%CI, 1.17-7.19; p<0.001; *I*^*2*^=100%; Figure 2, Panel A). Consistent results were found when sensitivity analyses were performed exclusively restricted to studies comparing patients requiring ICU admission versus no ICU admission (two studies; n=540; RoM=3.24; 95%CI, 2.54-4.14; p<0.001; *I*^*2*^=87%; Figure 2, Panel B) but not for the analysis of severe or critical scores versus mild (three studies; n=561; RoM=3.63; 95%CI, 0.65-20.37; p=0.14; *I*^*2*^=100%; Figure 2, Panel C). Statistical heterogeneity was elevated across all analyses and did not significantly improve with the planned sensitivity analyses.

**Figure 2.**
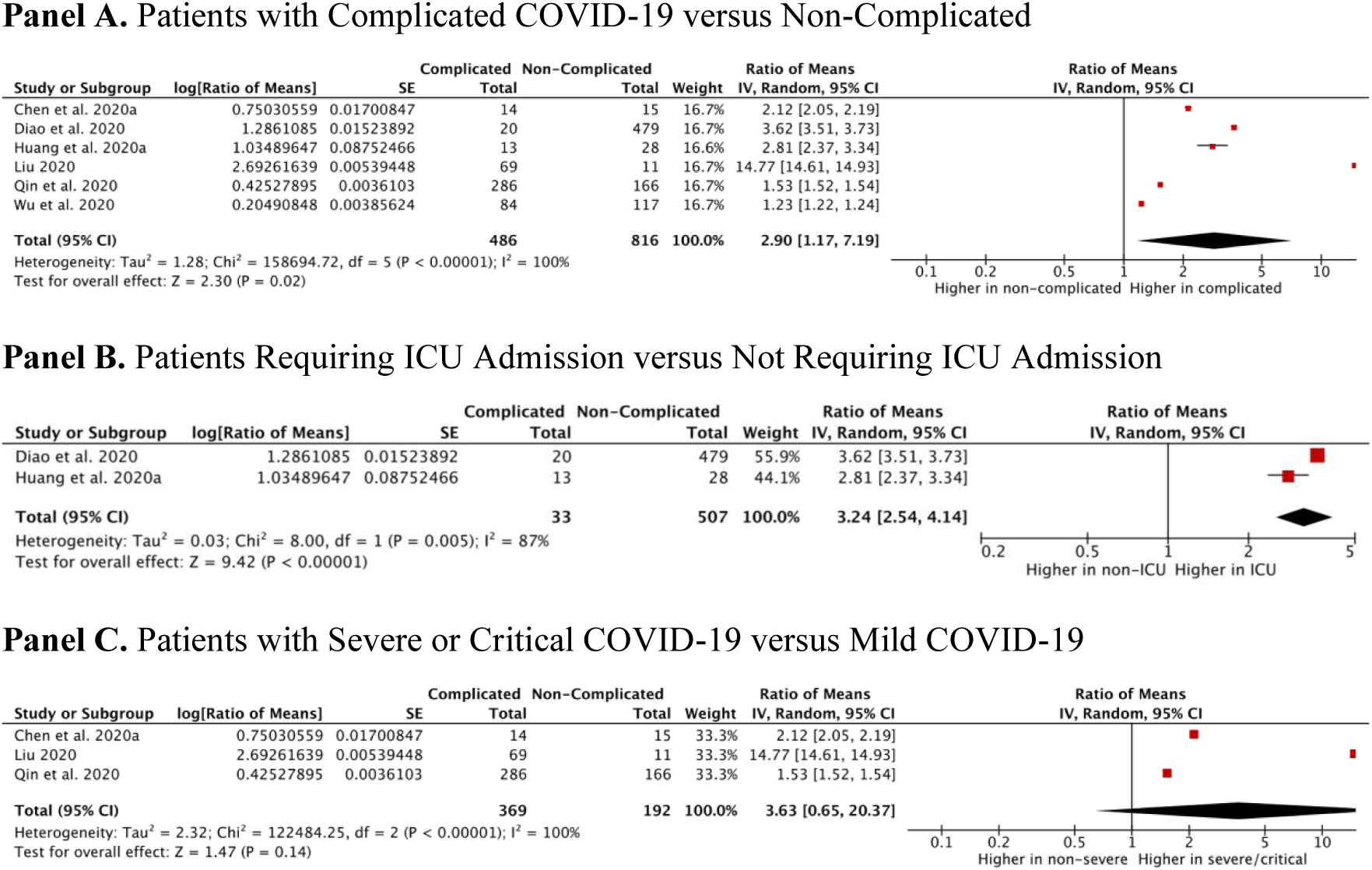
Meta-Analysis of Serum IL-6 Levels in COVID-19.

Baseline IL-6 positively correlated with bilateral pulmonary involvement (r=0.45, p=0.001), and maximum body temperature (r=0.52, p=0.001) in the retrospective cohort study by Liu et al.^37^ Among 30 patients with IL-6 assessment before and after treatment, 26 (87%) patients had significantly reduced IL-6 concordant with improving pulmonary computed tomography. In contrast, amongst the four patients who experienced progressive clinical deterioration, three (75%) had increasing IL-6 levels.

In the analysis of risk factors for ARDS and death by Wu et al.,^41^ patients with COVID-19 who progressed to ARDS had significantly increased IL-6 (median 7.39 pg/mL, IQR 5.63-10.89 versus median 6.29 pg/mL, IQR 5.36-7.83; p=0.03). Further, elevated IL-6 was associated with death. Similarly, Ruan et al.^39^ identified significantly higher IL-6 levels amongst patients who die from COVID-19 compared to those who survive (11.4±8.5 pg/mL versus 6.8±3.6 pg/mL, p<0.001).

### Tocilizumab and COVID-19

One study examined the clinical impact of tocilizumab on severe COVID-19 disease.^43^ In a non-randomized, open-label, clinical trial, 21 patients with severe or critical COVID-19 were treated with tocilizumab. Mean age was 56.8 years and 14% of patients were female; critical disease was identified in four patients (19%). All patients received institutional standard of care including lopinavir and methylprednisolone, as well as tocilizumab 400 mg intravenously in one to two doses. Eighteen patients (85.7%) received tocilizumab once, and three patients (14.3%) received tocilizumab twice, with the second dose being administered due to recurrence of fever within 12 hours of the first administration.

After receipt of tocilizumab, all patients had resolution of fever within 24 hours, with reported relief of clinical symptoms. There was a statistically significant decrease in oxygen requirements from day two to five after receipt of tocilizumab (15/20 (75%) of patients had reduced inhaled oxygen requirements). Further, there was resolution of radiographic abnormalities on chest computed tomography in 19 patients (90.5%). Mean CRP reduced from 75.06±66.80 mg/L pre-tocilizumab to 2.72±3.60 mg/L on day 5 post-tocilizumab. No patients died; 19 patients (90.5%) survived to discharge, and 2 remain admitted to hospital by the end of follow-up. No adverse drug reactions were identified, nor pulmonary infections after tocilizumab treatment.

Our search of clinical trial registries identified five registered studies investigating the therapeutic role of tocilizumab therapy in patients with COVID-19 (Table 3). Of these, all are based in China; one has since been cancelled by the principal investigator and one is planned but not yet recruiting. Two of the studies underway are multi-centre RCTs, with a third RCT pending recruitment. The cumulative anticipated sample size for the four planned studies will be n=518 and the methodological designs are complementary in target patient populations (all-comers, all-comers with elevated IL-6, severe cases with elevated IL-6, and severe cases with elevated IL-6 meeting diagnostic criteria for grade 2-3 CRS) and comparators (supportive therapy, favipiravir, continuous renal replacement therapy).

### Risk of Bias Assessment

In cohort studies assessing inflammatory response in COVID-19, risk of bias was assessed via the QUIPS tool.^30,31^ Four studies were determined to be at high risk of bias,^37,39,41,42^ five moderate,^33,35,36,38,40^ and one low (Supplementary Figure 1);^34^ this was mostly driven by lack of control for confounding and potential inconsistencies in the measurement of the inflammatory mediators under study. For the assessment of tocilizumab treatment for COVID-19,^43^ risk of bias was determined to be severe following assessment by the ROBINS-I tool.^32^

## DISCUSSION

In this systematic review and meta-analysis, we demonstrate that serum levels of IL-6 are significantly elevated in the setting of severe COVID-19 disease. Meta-analysis of the available data indicates that such increased levels are significantly associated with adverse clinical outcomes, including ICU admission, ARDS, and death. Patients with such complicated forms of COVID-19 had nearly three-fold higher serum IL-6 levels than those with non-complicated disease. In the single, non-randomized study published investigating the effect of tocilizumab treatment, decreased levels of serum IL-6, improved clinical and radiological status, and the absence of mortality were observed.

It is increasingly recognized that a dysregulated host immune response to foreign infectious pathogens is integral to the development of target organ dysfunction and a major contributor to morbidity and mortality. Specifically, the systemic inflammatory response in sepsis has been demonstrated to overlap with that of CRS;^44,45^ in patients with COVID-19 complicated by ARDS, such a hyperactivation of the humoral immune system with a prominent IL-6 response may suggest that part of the pathogenesis of complicated disease involves a dysregulated and excessive host inflammatory response. This clinical phenotype resembles that of CRS, a condition for which IL-6 receptor inhibition with tocilizumab has clearly demonstrated benefit^21^ and may represent a more severely affected COVID-19 subpopulation, with increased requirements for critical care and worse clinical outcomes.^47^

The results of the preliminary investigation of tocilizumab in severe COVID-19 are promising, but also suffer from important limitations.^43^ In patients receiving tocilizumab, all experienced rapid resolution of fever, symptomatic improvement, and the majority had early reductions in oxygen requirements and radiographic abnormalities. In contrast to other cohorts, which have demonstrated mortality rates of up to 61.5% amongst critically ill patients (and the pooled mortality of 22% among all hospitalized patients seen in our meta-analysis), no patients died in this study after administration of tocilizumab.^48^ Further, despite intensive immunosuppression, no patients developed superimposed nosocomial pulmonary infections nor significant adverse drug reactions. This study is limited by its small sample size, non-randomised intervention, lack of comparator arm, and retrospective design, but the results are biologically plausible as the included patients all had elevated IL-6 levels which decreased progressively in parallel to clinical and radiographical improvement.^43^

Despite these early signals of benefit, the results of several ongoing clinical studies should be awaited to better define the role of tocilizumab in the therapy of COVID-19.^49-53^ Such studies benefit from larger sample sizes, prospective recruitment of patients with a wide spectrum of disease severity, and in some protocols, the use of active controls. Further, sarilumab, another monoclonal antibody blocking the IL-6 receptor, is being planned for a United States-based adaptive multi-centre RCT in severe COVID-19 infection, but is not yet registered.^54^

### Limitations

Although designed and reported in accordance with standardized systematic review methodology^25,26^ and employing a highly sensitive search strategy, including the grey literature, this study has important limitations, much of which is inherent to the methodological quality of the included primary studies. All primary studies eligible for inclusion were conducted in China, with several studies recruiting participants from the same centres; while none of the included studies described their data as having been previously published, this remains a theoretical possibility.^55^

We encountered high levels of statistical heterogeneity in our meta-analysis comparing IL-6 levels between patients with complicated and non-complicated disease; although we performed pre-specified sensitivity analyses, these failed to sufficiently explain this heterogeneity. Such residual heterogeneity may have arisen from multiple sources of variability between studies, most prominently due to likely differences in patient characteristics, lack of consecutive enrolment, variable timing of IL-6 measurement, the absence of a set definition of “supportive care”, and differences in adjuvant immunomodulatory medications received, such as corticosteroids and IVIg, which may have affected both IL-6 response and patient outcomes.

Most studies included in this review were rated at moderate or high risk of bias, reflecting generally low methodological quality. This was primarily driven by a lack of control for confounding, inconsistencies or unclarity of the context in which IL-6 measurements were performed, and potential for selection bias due to lack of consecutive patient enrolment. Lastly, we identified only a single, non-randomized, single-arm study investigating the effect of tocilizumab treatment in patients with COVID-19; while promising, this provides only low-level evidence for the efficacy and safety of this drug for the treatment of severe COVID-19 and should be primarily considered hypothesis-generating at this time.

## CONCLUSIONS

In this systematic review and meta-analysis, we demonstrate serum levels of IL-6 to be significantly elevated in the setting of complicated COVID-19 disease, and increased IL-6 levels to be in turn significantly associated with adverse clinical outcomes. This suggests that the progression of COVID-19 to complicated disease may be the consequence of an excessive host immune response and autoimmune injury. Preliminary investigations indicate that the inhibition of the cytokine pathway at the level of IL-6 with tocilizumab may be efficacious in managing this dysregulation. Whilst preliminary in nature, this data identifying increased IL-6 levels in severe COVID-19 disease and early signals of clinical benefit for tocilizumab therapy support the need for ongoing clinical studies to elucidate the role of IL-6 inhibition in the therapy of severe COVID-19.

## Data Availability

This study meta-analyzes study-level data from peer-reviewed, published research and cites these sources within the manuscript. The data can thus be accessed from the cited studies.

---

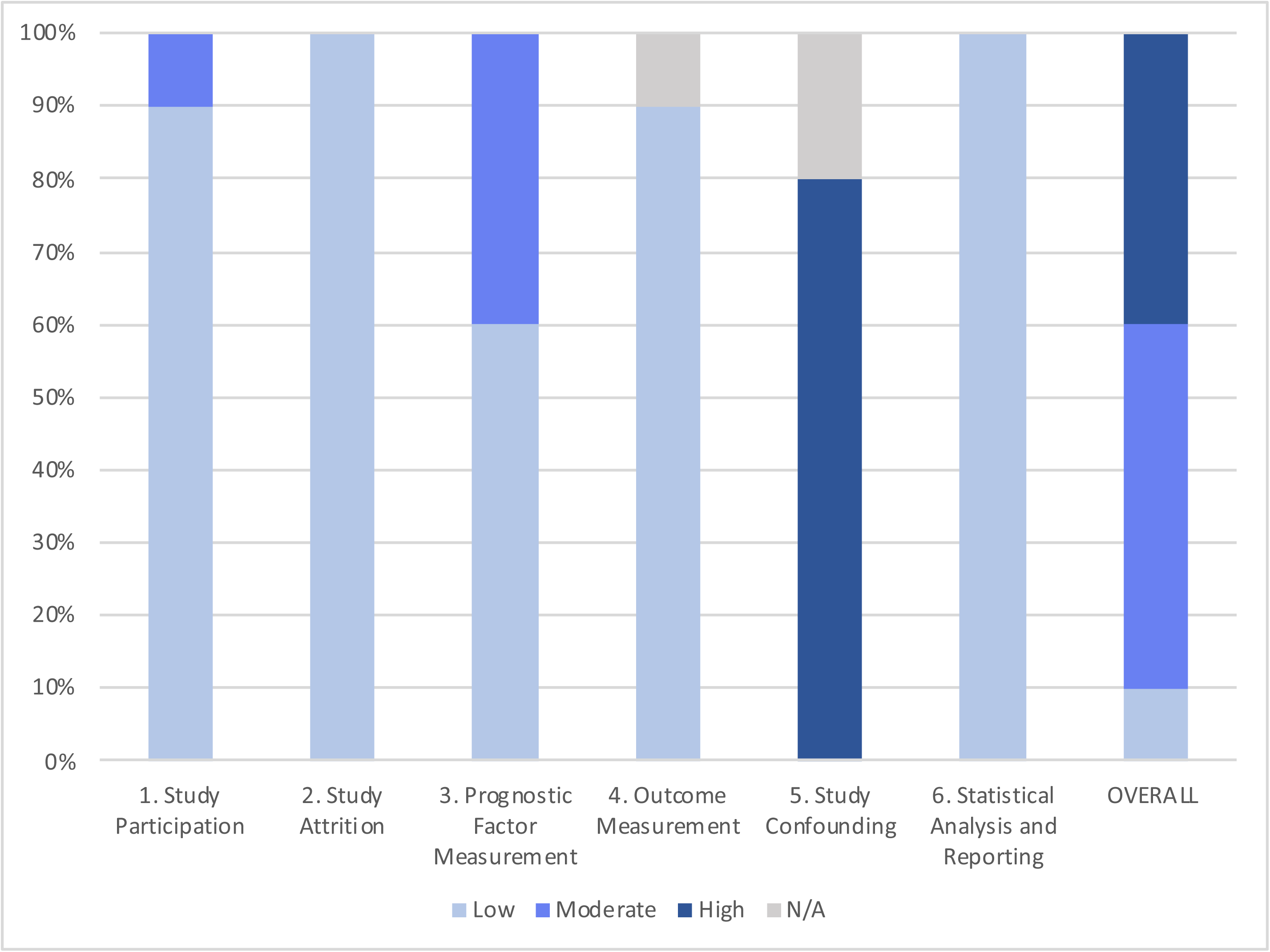

## Notes

### Competing Interest Statement

The authors have declared no competing interest.

### Clinical Trial

PROSPERO Registration – CRD42020175879

### Clinical Protocols

https://www.crd.york.ac.uk/prospero/display_record.php?ID=CRD42020175879

